# Development and International Validation of a Deep Learning Model for Predicting Acute Pancreatitis Severity from CT Scans

**DOI:** 10.1101/2025.07.06.25330963

**Authors:** Yanqi Xu, Brigitta Teutsch, Weicheng Zeng, Yang Hu, Shikhar Rastogi, Emmy Yuebi Hu, Isabella DeGregorio, Wan Fung Chui, Benjamin I. Richter, Ryan Cummings, Julia E. Goldberg, Edwin Mathieu, Belinda Appiah Asare, Péter Hegedűs, Kriszta-Beáta Gurza, István Viktor Szabó, Hedvig Tarján, Andrea Szentesi, Ruben Borbély, Dorottya Molnár, Nándor Faluhelyi, Áron Vincze, Katalin Márta, Péter Hegyi, Qi Lei, Tamas Gonda, Chenchan Huang, Yiqiu Shen

## Abstract

**Background and aims:** Acute pancreatitis (AP) is a common gastrointestinal disease with rising global incidence. While most cases are mild, severe AP (SAP) carries high mortality. Early and accurate severity prediction is crucial for optimal management. However, existing severity prediction models, such as BISAP and mCTSI, have modest accuracy and often rely on data unavailable at admission. This study proposes a deep learning (DL) model to predict AP severity using abdominal contrast-enhanced CT (CECT) scans acquired within 24 hours of admission.

**Methods:** We collected 10,130 studies from 8,335 patients across a multi-site U.S. health system. The model was trained in two stages: (1) self-supervised pretraining on large-scale unlabeled CT studies and (2) fine-tuning on 550 labeled studies. Performance was evaluated against mCTSI and BISAP on a hold-out internal test set (n=100 patients) and externally validated on a Hungarian AP registry (n=518 patients).

**Results:** On the internal test set, the model achieved AUROCs of 0.888 (95% CI: 0.800–0.960) for SAP and 0.888 (95% CI: 0.819–0.946) for mild AP (MAP), outperforming mCTSI (p = 0.002). External validation showed robust AUROCs of 0.887 (95% CI: 0.825–0.941) for SAP and 0.858 (95% CI: 0.826–0.888) for MAP, surpassing mCTSI (p = 0.024) and BISAP (p = 0.002). Retrospective simulation suggested the model’s potential to support admission triage and serve as a second reader during CECT interpretation.

**Conclusions:** The proposed DL model outperformed standard scoring systems for AP severity prediction, generalized well to external data, and shows promise for providing early clinical decision support and improving resource allocation.

## Introduction

Acute pancreatitis (AP) is a common gastrointestinal disease with a rising global incidence.^1–3^ In the United States, it ranks among the top causes of gastrointestinal-related hospitalizations, accounting for 90 admissions per 100,000 hospital visits.^4^ While over 80% of AP are mild and self-limiting, about 5% progress to severe AP, which carries a mortality rate as high as 30-40%.^5,6^ A major challenge in AP management is that current severity classification, as defined by the Revised Atlanta Classification (RAC)^7^, depends on the development of clinical complications that typically emerge 48–72 hours after symptom onset, which delays effective intervention.^8^ Accurate severity prediction at the time of admission is essential for improving care: enabling timely discharge of mild AP patients, while preparing intensive care unit (ICU) admission for those that will progress to severe AP, particularly in resource-constrained settings.

Laboratory biomarkers, such as C-reactive protein (CRP), blood urea nitrogen (BUN), and hematocrit have been explored for predicting AP severity. However, CRP is limited for early triage as it peaks at 48–72 hours after symptom onset,^9^ while BUN and hematocrit mainly reflect volume status rather than pancreatic injury, making them prone to confounding from dehydration, renal dysfunction, or anemia.^10^ As a result, multi-factorial models such as APACHE-II,^11^ Ranson Score,^12^ Glasgow,^13^ EASY-APP,^14^ and Bedside Index of Severity of Acute Pancreatitis (BISAP)^15^ remain the most commonly used tools for early severity prediction. These systems integrate both clinical and laboratory data but have only moderate accuracy with significant variability across cohorts, with reported areas under receiver operating curve (AUROCs) ranging from 0.57 to 0.84.^16–19^ Moreover, many of these high-performing systems such as APACHE-II require a large number of parameters or depend on clinical or laboratory values that take over 48 hours to collect, limiting their utility in real-time decision-making.^9,20,21^

Imaging-based scoring systems have been developed, predominantly using contrast-enhanced computed tomography (CECT). The Modified CT Severity Index (mCTSI) is the most widely used imaging-based prognostic method, which requires visual assessment of pancreatic inflammation, necrosis, and extrapancreatic complications from CECT images.^22^ However, studies indicate that mCTSI offers no clear advantage over clinical scoring systems.^23,24^ Furthermore, its reliability is undermined by high inter-observer variability.^25–27^ In addition, key mCTSI markers, such as pancreatic necrosis, could remain radiologically undetectable until 72-96 hours after symptom onset, limiting its utility for early severity prediction.^28–30^

Despite these limitations, the majority of patients presenting to the emergency room in the U.S. with abdominal pain will undergo abdominal CT exam.^31^ This makes CT imaging a valuable resource for developing deep learning (DL) models to predict AP severity. Convolutional neural networks (CNNs) trained on CECT and non-contrast CT have demonstrated improved accuracy over traditional scoring systems.^32–34^ However, most of these models are trained on single-center datasets without external validation, raising concerns about their generalizability. Furthermore, existing methods typically rely on manually annotated severity categorization based on the RAC guidelines as a reference standard, which are difficult to obtain at large-scale.

To address these limitations, we developed and validated a DL model to predict AP severity using CECT scans obtained at admission, focusing on the accurate identification of mild acute pancreatitis (MAP) and severe acute pancreatitis (SAP). These two extremes are clinically important: MAP patients may be safely discharged early, while SAP patients require intensive monitoring and timely escalation of care. To reduce reliance on manual annotations, we employed self-supervised learning (SSL) to pretrain the model on large-scale unlabeled CT data, followed by fine-tuning on a curated labeled dataset. We evaluated the model’s performance against two widely used clinical scoring systems, mCTSI^22^ and BISAP^15^, on both an internal test set collected in the U.S. and an external, prospectively maintained Hungarian registry encompassing a heterogeneous, multi-center patient population. BISAP was chosen as a representative clinical-variable-based model, and mCTSI as a commonly used imaging-based model, together reflecting the current standard of care. To further explore clinical utility, we conducted retrospective simulations of risk-based triage. An overview of the study design is provided in Figure 1.

**Figure 1.**
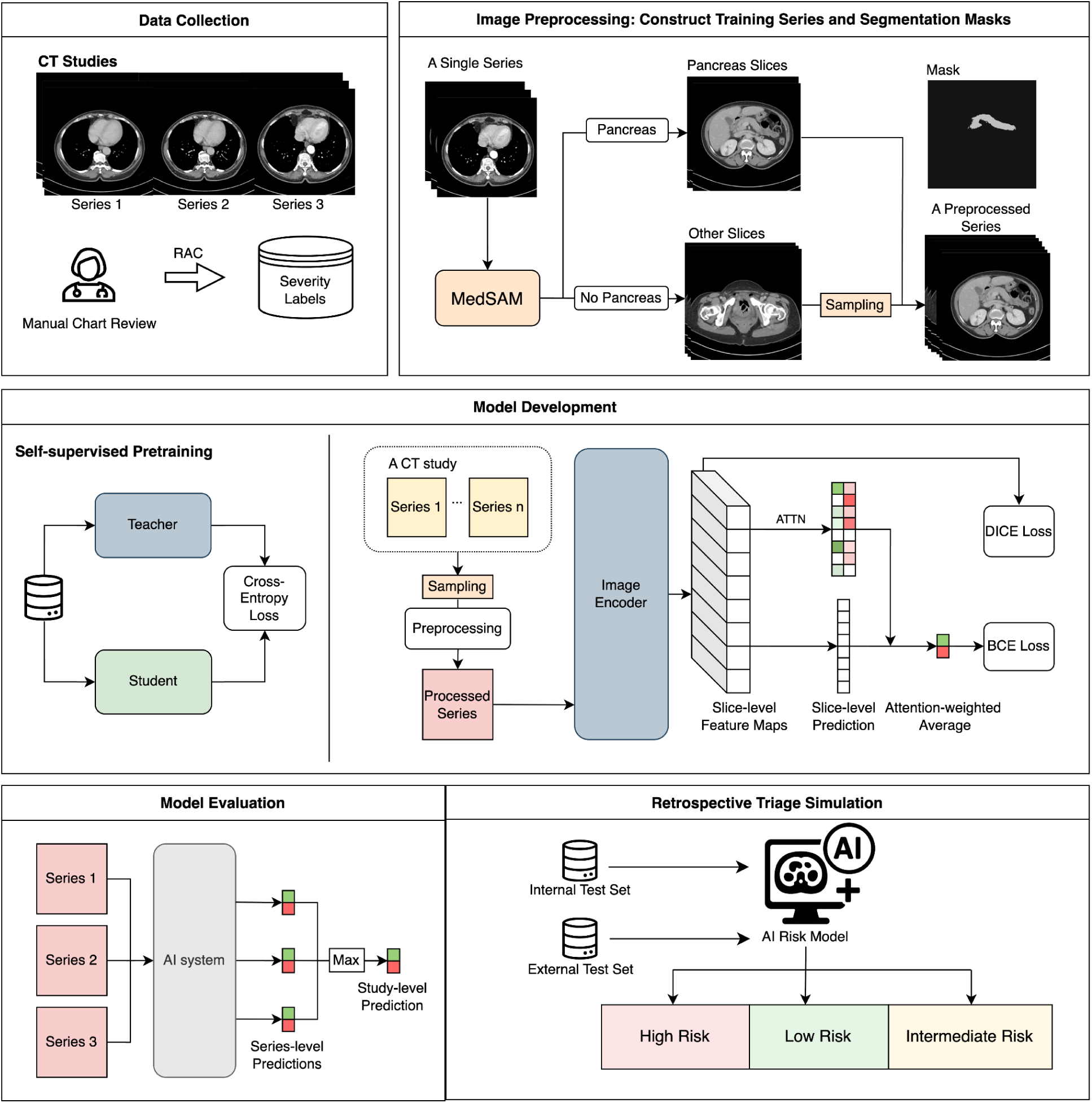
Workflow of the AI system. Data Collection: CT studies with multiple series were used in this study. Severity labels were assigned based on the Revised Atlanta Classification (RAC). **Image Preprocessing**: During training, MedSAM, an automated deep learning segmentation model, was used to generate pancreas masks and identify relevant slices. These were combined with randomly sampled non-pancreas slices to create preprocessed input series. Segmentation is not required at inference time. **Model Development:** A transformer-based backbone (pretrained via self-supervised learning) extracted slice-level features. Slice scores derived from saliency maps were aggregated using attention (ATTN) to generate severity predictions, optimized with binary cross entropy (BCE), Dice, and L1 losses. **Model Evaluation:** Study-level predictions were generated by using the maximum series-level score. **Retrospective Triage Simulation** was performed on both internal and external test sets.

## Methods

This retrospective study was approved by the NYU Langone Institutional Review Board (IRB Protocol #I24-00008) and the requirement for informed consent was waived. All procedures adhered to the ethical principles of the Declaration of Helsinki. The study design and reporting comply with the CLAIM^35^ and TRIPOD guidelines.^36^

### Data Collection

The data collection and filtering pipeline for the development cohort, internal test set, and external test set is illustrated in Figure 2. All scans were stored in DICOM format, retrieved from the institutional PACS and de-identified by removing all protected health information (PHI) from DICOM metadata. Details of the image acquisition protocol are provided in Supplementary Materials S1.1, and the severity labeling and mCTSI scoring procedure are described in Supplementary Materials S1.2.

**Figure 2.**
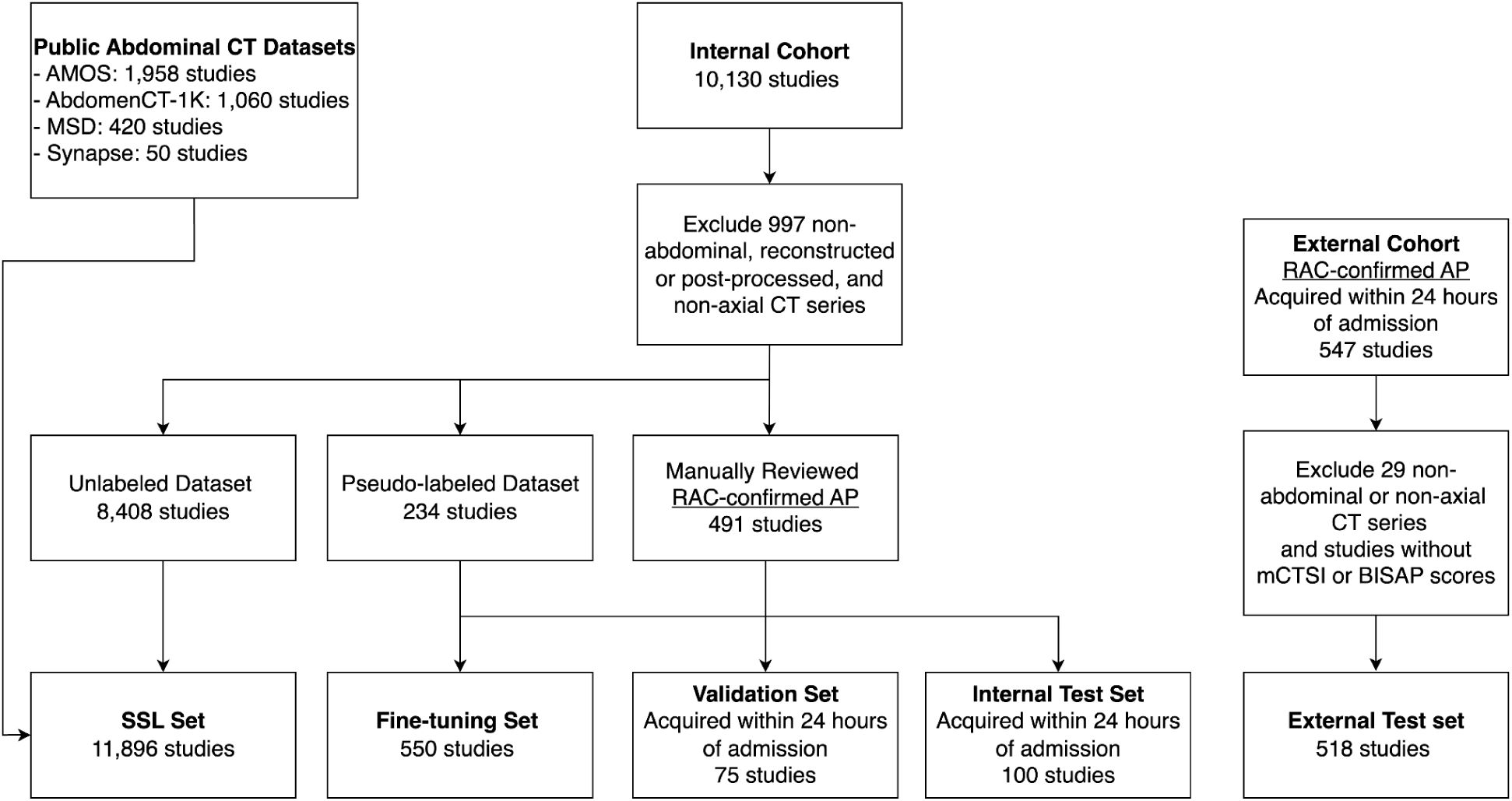
Data Cleaning and Modeling Workflow. Non-abdominal CT series, reconstructed/post-processed images, and non-axial views were excluded from both internal and external cohorts. In total, 491 studies in the internal cohort and all 547 studies in the external cohorts were manually reviewed, with acute pancreatitis diagnosis confirmed per Revised Atlanta Classification (RAC) criteria. The DL model was pre-trained using self-supervised learning (SSL) on 8,408 unlabeled CT studies from the internal cohort and 3,488 studies from four public abdominal CT datasets. Fine-tuning used 550 studies: 234 manually labeled (RAC criteria) and 316 pseudo-labeled from radiology reports (details in Supplementary Materials S2). Evaluation was performed on an internal test set of 100 studies and an external test set of 518 studies, both acquired within 24 hours of admission and manually annotated using RAC criteria.

### Development and Internal Test Sets

We collected 10,130 CECT studies from 8,335 patients at NYU Langone Health between November 2002 and January 2025. NYU Langone Health is a large integrated academic health system that encompasses major tertiary urban medical centers, as well as suburban hospital centers, and a broad network of ambulatory care sites.

Eligibility criteria included age ≥ 18 years and a radiology report confirming the diagnosis of AP. Of these studies, 491 were reviewed to confirm the presence of abdominal pain, absence of pancreatic malignancy, and an AP diagnosis meeting RAC criteria.^7^ AP severity scoring for these studies were retrospectively labeled according to the RAC^7^, based on both CT imaging and electronic health record (EHR) review. The mCTSI scores were independently assessed by senior radiology trainees.

The 491 manually labeled studies were randomly split into 316 for fine-tuning, 75 for validation, and 100 for internal testing. The split is performed at the patient level to prevent data leakage.

Of note, all CT studies in the validation and test sets were acquired within 24 hours of the patients’ initial hospital admission. To expand the fine-tuning set, 234 additional AP severity labels were extracted from radiology reports using rule-based string-matching. We refer to these as pseudo-labels and these pseudo-labeled studies were used exclusively for model fine-tuning (details in Supplementary Materials S2). The final fine-tuning cohort thus comprised 550 labeled studies (316 manually labeled and 234 pseudo-labeled).

To leverage the power of large-scale unlabeled data, we constructed a SSL dataset comprising 11,896 unlabeled CT studies. This dataset was created by combining the remaining unlabeled 8,408 CT studies from the development cohort with four public abdominal CT datasets: AMOS^37^ (n=1,958), AbdomenCT-1K^38^ (n=1,060), MSD^39^ (n=420), and Synapse^40^ (n=50).

In total, our development cohort comprised 12,521 CT studies, including 11,896 unlabeled studies used for SSL pre-training, 550 pseudo- or manually labeled studies for fine-tuning, and 75 manually labeled studies for validation. The internal test set included 100 manually labeled studies that were entirely independent of the development cohort.

#### External Test Dataset

The external test dataset was obtained from the GOULASH trial,^41^ a multicenter, randomized controlled study including patients from three tertiary medical centers in Hungary (Pécs, Székesfehérvár, Budapest) between 2017 and 2023.^42^ Inclusion criteria were the same as for the internal test set: age over 18, presence of abdominal pain, AP diagnosis based on the RAC,^7^ no history of pancreatic malignancy, and availability of CT imaging within 24 hours of admission. A total of 619 patients were enrolled in the study, from whom 547 abdominal CT studies were retrieved at participating centers, anonymized and transferred to NYU via Globus for analysis. This dataset reflects real-world heterogeneity, with imaging acquired using various scanner vendors, models, and protocols. We excluded non-abdominal series, non-axial images, and studies lacking valid mCTSI or BISAP scores, resulting in a final dataset of 518 CT studies. AP severity assessment was carried out at the end of hospitalization by a multidisciplinary team, including gastroenterology residents and specialists, based on the RAC^7^. The mCTSI score was retrospectively independently assessed by six radiology residents under attending radiologist supervision, all of whom were blinded to the clinical information.

### Image Preprocessing

Each abdominal CT study comprises multiple series, which are groups of slices acquired with consistent scan settings and uniquely identified by the SeriesInstanceUID in the DICOM metadata. We excluded non-abdominal series, series with post-processed and reconstructed images (e.g. BONE reconstructed), and non-axial series (sagittal and coronal planes). Figure 2 outlines the exclusion criteria. All images are converted to Hounsfield unit (HU) and windowed with a width of 400 and center of 45. Each 2D axial slice was resized to 256 by 256 for model training and inference.

#### Identifying pancreas-related slices

Only a subset of slices in an abdominal CT study captures the pancreas. During training, to focus the model on the most relevant anatomy, we used MedSAM^43^, an off-the-shelf segmentation model, to generate a binary pancreas mask for each slice. We computed the sum of pixel values within each segmentation mask to quantify the presence of pancreatic tissue. The 64 consecutive slices with the highest average mask sum within a single series are selected, as they are most likely to contain the pancreas in its largest cross-sectional regions. Remaining slices were considered non-pancreatic. Of note, slice selection is only applied during training. At test time, the model processed all available slices from each CT study without requiring pancreas slice selection. Details of the segmentation process are provided in Supplementary Section S1.3.

### Problem Formulation

We framed AP severity prediction as an ordinal classification problem^44^ and decomposed it into two binary classification tasks:

- MAP versus non-MAP,
- SAP versus non-SAP.

Severity labels were encoded as follows: MAP cases were assigned [0, 0]; Moderately severe AP (MSAP) cases were assigned [1, 0]; SAP cases were assigned [1, 1]. The model outputs two probabilities [𝑝_0_, 𝑝_1_], where 𝑝_0_ is the probability of MAP, and 𝑝_1_ is the conditional probability of SAP given non-MAP. Final severity probabilities are computed as:

- Probability of MAP: 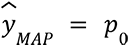
- Probability of SAP: 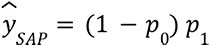

This two-step approach preserved the ordinal nature of AP severity by ensuring that predictions reflect the clinical progression from mild to moderately severe to severe.

### Deep Learning Model Architecture

The model workflow is illustrated in Figure 1. The input consists of a series of CT slices from each patient, each independently processed by a Vision Transformer (ViT)^45^ base encoder to generate a slice-level feature map. A 1x1 convolution is then applied to produce a 2D saliency map for each feature map, which highlights regions most relevant for severity prediction. To summarize slice-level information, we compute a scalar slice score by aggregating the top 𝑘% of the pixel values from the saliency map. The resulting set of slice scores is passed to an attention-based aggregation head,^46^ which generate two final severity predictions 𝑝_0_ and 𝑝_1_, using attention-weighted averaging. To compute attention weights, each slice-level feature map undergoes global average pooling to produce a feature vector. These vectors are then passed through two independent attention mechanisms,^46^ each generating task-specific weights that determine the contribution of each slice to the final prediction. This allows the model to focus on the most informative slices for each classification task.

### Pre-training and Fine-tuning

To leverage large-scale unlabeled data, we pre-trained our image encoder using DINO^47^ and DINOv2^48^ on 11,896 unlabeled pancreas CT studies. DINO and DINOv2 train the image encoder to extract consistent feature representations across different augmented views of the same image. Additional details can be found in Supplementary Materials S4. During fine-tuning, each input series was constructed by selecting the 64 slices containing the pancreas (as defined by segmentation masks), and *M* (treated as a hyperparameter) additional slices sampled from the remaining non-pancreas slices. One series was randomly selected and a new set of non-pancreas slices was randomly sampled from each CT study at each epoch. During evaluation, the model processed all available slices from each CT study without requiring pancreas slice selection. A prediction was generated for each series, and the final study-level prediction was taken as the maximum score across all series, assuming the most informative series reflected the patient’s true severity. This design simplified deployment by eliminating the need for segmentation. Additional training details, including loss functions, hyperparameters choices (e.g. 𝑘 and 𝑀) and data augmentations are in Supplementary Materials S3.

### Operating Point Selection

Thresholds were selected to convert mCTSI, BISAP and AI risk scores into binary decisions. For mCTSI, established cutoffs were used: ≤2 for low-risk (MAP) and ≥8 for high-risk (SAP).^22^ A BISAP score of ≥3 is considered as high-risk and ≤1 for low-risk.^49^ For performance comparisons on the internal and external test sets, AI thresholds were selected to match the specificity of mCTSI. For the risk-based triage simulation, AI thresholds, 𝑡*_M_* for low-risk and 𝑡*_S_* for high-risk, were selected from the internal validation set to jointly optimize sensitivity and positive predictive value (PPV), with the goal of surpassing mCTSI performance.

### Model Evaluation and Statistical Analysis

Model performance was evaluated using area under the receiver operating characteristic curve (AUROC), area under the precision-recall curve (AUPRC), sensitivity, specificity, positive predictive value (PPV), and negative predictive value (NPV), with 95% confidence intervals (CIs) computed via 10,000 bootstrap samples. Results are based on an ensemble of the top five models with the highest AUROC on the validation set of the development cohort. AUROC comparisons between the AI model, mCTSI, and BISAP were conducted using the DeLong test^50^; subgroup comparisons used unpaired DeLong tests.^51^ Sensitivity and specificity were compared using permutation test. All metrics were computed using the scikit-learn package (v1.6.1)^52^, and DeLong tests were executed using the pROC package^51^ (v1.18.5) in R (v4.5.1).

## Results

The internal test set included 100 abdominal CT studies from 100 patients (41% female; mean age: 54.33 ± 18.01 years), with severity labels distributed as follows: 40 MAP (40%), 48 MSAP (48%), and 12 SAP (12%). None of these patients overlapped with the development cohort. The external test set comprised 518 abdominal CT studies from 518 patients (40.93% female; mean age: 54.42 ± 14.83 years), with 276 MAP (53.3%), 209 MSAP (40.4%), and 33 SAP (6.4%). This external cohort was entirely independent of the development datasets.

### Performance on the Internal Test Set

For identifying high-risk patients (SAP versus non-SAP), the AI model achieved an AUROC of 0.888 (95% CI: 0.800–0.960), outperforming mCTSI (AUROC 0.704; 95% CI: 0.556–0.839; p = 0.002). While mCTSI showed high specificity (0.92; 95% CI: 0.859–0.976), its sensitivity was limited (0.25; 95% CI: 0.0–0.5). At the same specificity level, the AI model achieved a sensitivity of 0.50 (95% CI: 0.200–0.800, p=0.123). For identifying low-risk patients (MAP vs. non-MAP), the AI model achieved an AUROC of 0.888 (95% CI: 0.819–0.946), outperforming mCTSI (AUROC: 0.816, 95% CI: 0.733–0.890; p < 0.001). At the matched specificity (0.850; 95% CI: 0.755–0.933), the AI model achieved a sensitivity of 0.75 (95% CI: 0.610–0.878) compared to mCTSI (0.675, 95% CI: 0.525–0.818, p=0.291). Model performance on the internal test set is summarized in Table S5.1 and Figure 3(A). Additional metrics, including AUPRC, sensitivity, specificity, PPV, and NPV, are reported in Table S5.1.

**Figure 3.**
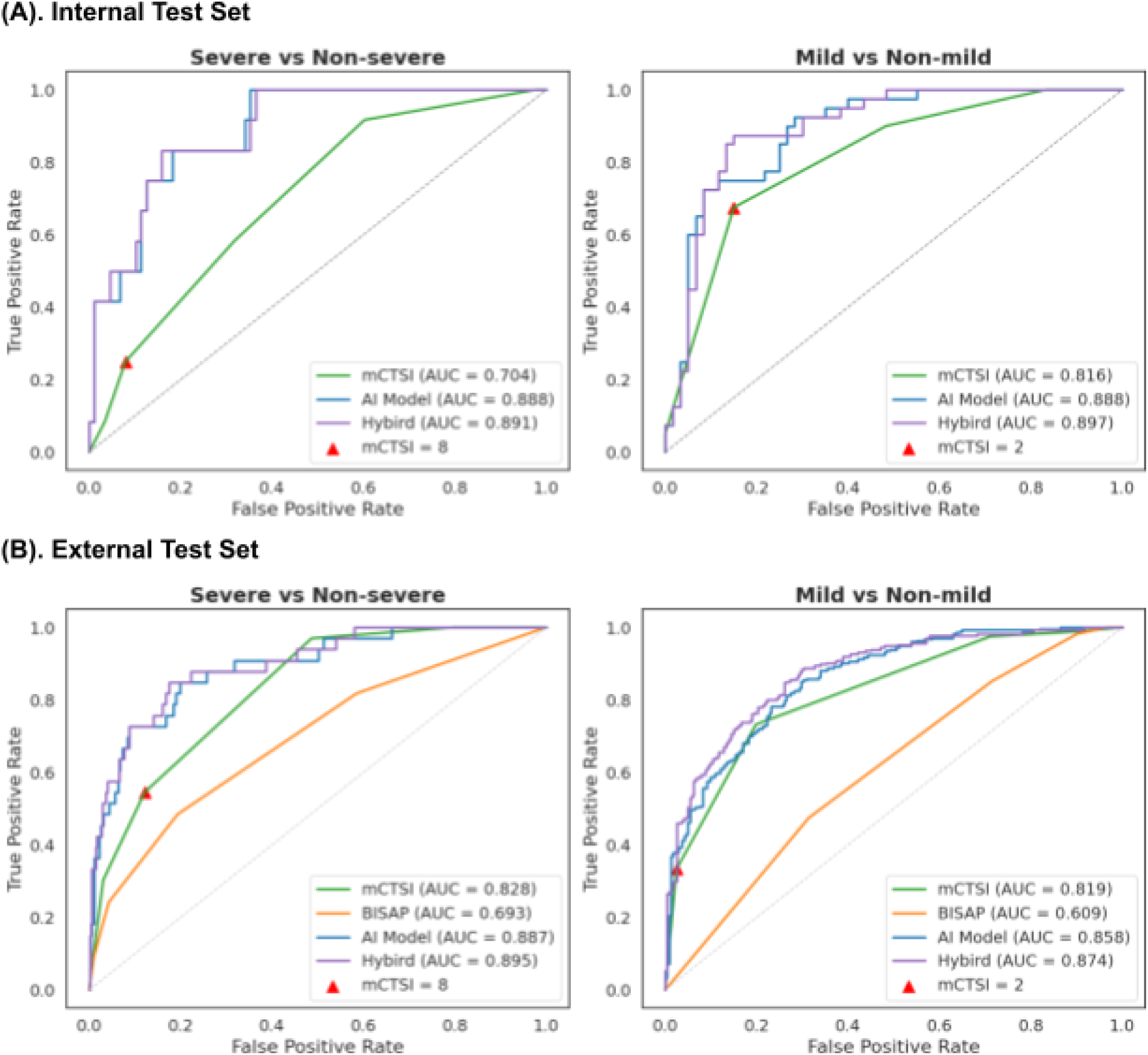
ROC Curves for the AI Model, mCTSI, BISAP, and Hybrid Model on (A) Internal (n = 100 patients) and (B) External (n = 518 patients) Test Sets. Left panels show ROC curves for identifying severe acute pancreatitis (SAP); right panels show ROC curves for identifying mild acute pancreatitis (MAP). Operating points used for mCTSI risk stratification are indicated on the curves. On both datasets and across both tasks, the AI model outperformed mCTSI, and on the external test set, it also outperformed BISAP. The hybrid model that combined mCTSI and AI predictions further improved performance.

### Performance on the External Test Set

The AI model maintained strong performance on the external test set, showing no degradation compared to the internal evaluation. For identifying high-risk cases (SAP vs. non-SAP), the AI model achieved an AUROC of 0.887 (95% CI: 0.825–0.941), outperforming mCTSI (0.828, 95% CI: 0.769–0.883; p = 0.024) and BISAP (0.693, 95% CI: 0.592–0.787; p = 0.002). Similarly, for identifying low-risk patients (MAP vs. non-MAP), the model achieved an AUROC of 0.858 (95% CI: 0.826–0.888), surpassing mCTSI (0.819; 95% CI: 0.785–0.851; p = 0.019) and BISAP (0.609; 95% CI: 0.564–0.654; p < 0.001). Model performance on the external test set is summarized in Table S5.2 and Figure 3(B). Additional metrics are reported in Table S5.2.

### Impact of Self-Supervised Learning and Pseudo-Labeling

We conducted ablation studies on the external test set to evaluate the impact of SSL pre-training and pseudo-labeled data on model performance. The baseline model, trained without SSL pre-training and without pseudo-labeled data, achieved an AUROC of 0.799 (95% CI:0.717–0.871) for identifying SAP cases. Incorporating pseudo-labeled data increased the AUROC to 0.845 (95% CI: 0.770–0.907, p < 0.001), while using SSL-pretrained weights improved it to 0.846 (95% CI: 0.773–0.906, p = 0.007). Combining both approaches produced the best performance, with an AUROC of 0.887 (95% CI: 0.825–0.941). Similar trends were observed identifying MAP cases. Results are summarized in Supplementary Table S4. These findings demonstrate that SSL pre-training and pseudo-labeling improve model performance while reducing reliance on labor-intensive manual annotations.

### Subgroup Analyses

We conducted subgroup analyses to evaluate the AI model’s performance across age and gender in both the internal and the external test sets. As different age thresholds have been explored in AP severity scoring systems,^53^ we selected 55 years as the cutoff to ensure balanced subgroup sizes. For identifying low-risk cases, the model showed modest differences in performance across both gender (male vs. female) (internal: 0.881 vs 0.911, p = 0.615; external: 0.853 vs 0.873, p = 0.612) and age (≤55 vs. >55) (internal: 0.862 vs 0.906, p = 0.484; external: 0.867 vs 0.851, p = 0.538) subgroups in both datasets, but these differences were not statistically significant. Similarly, the model demonstrated statistically insignificant difference in performance across all subgroups for identifying high-risk cases prediction in the internal test set (gender: 0.937 vs 0.887, p = 0.677; age: 0.886 vs 0.914, p = 0.619). In the external test set, AUROCs for gender subgroups ranged from 0.840 to 0.939, showing some variability, although these differences were not statistically significant (p = 0.084). For age subgroups, AUROC values ranged from 0.870 to 0.930, with no significant differences observed (p = 0.237). The results are presented in Supplementary Figure S6 and Supplementary Table S6.

### Risk-Based Triage Simulation

To evaluate the AI model’s clinical utility in triaging AP patients, we conducted a retrospective simulation (Figure 4), that stratified patients into three risk categories based on model predictions:

● High risk: patients with AI-predicted probability of SAP above threshold 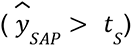 were classified as high-risk and considered for ICU admission.
● Low risk: patients with AI-predicted probability of SAP below or equal to the threshold (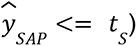 and AI-predicted probability of MAP above threshold 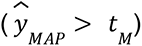 were classified as low-risk, suggesting eligibility for early discharge and conservative management.
● Intermediate risk: remaining patients were labeled intermediate-risk, warranting standard inpatient monitoring and evaluation.

**Figure 4.**
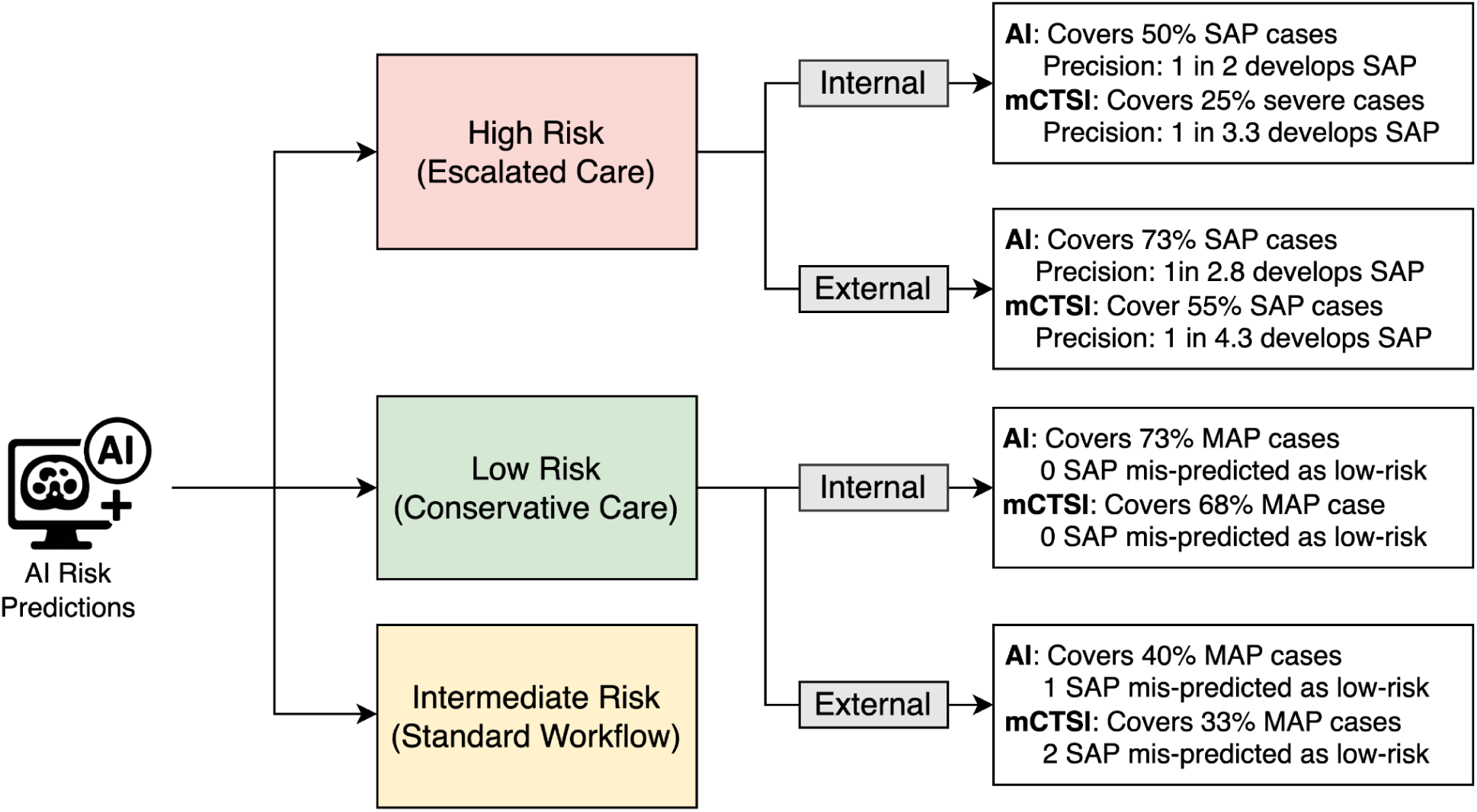
Clinical Simulation of Risk Stratification Using AI and mCTSI Predictions. Patients were categorized into low-risk, intermediate-risk, and high-risk groups based on risk scores from the AI model and mCTSI. Thresholds are described in the Method section ‘Operating Point Selection’. On the internal test set (n=100 patients), the AI model correctly identified 73% of mild acute pancreatitis (MAP) patients as low risk without misclassifying any severe cases. On the external test set (n=518 patients), the AI model identified 73% of severe acute pancreatitis (SAP) patients as high risk, with a precision of 1 severe case per 2.8 high-risk cases. These results highlight the AI model’s potential to improve risk stratification and support clinical decision-making.

Thresholds were determined using the method described in the ’Operating Point Selection’ section.

In the internal test set, the AI model stratified 12% (95% CI: 6%-19%) of patients as high-risk and 34% (95% CI: 25%-43%) as low-risk. The AI model successfully identified 50% (95% CI: 21%-80%) of SAP cases in the entire internal test set as high-risk, compared to mCTSI which identified 25% (95% CI: 0%-50%, p = 0.12). Among those classified as high-risk by the AI model, 1 in 2 patients (PPV: 0.5, 95% CI: 0.2-0.8) had SAP, compared to 1 in 3.3 for mCTSI (PPV: 0.3, 95% CI: 0.0-0.625, p = 0.11). For low-risk triage, the AI model identified 73% (95% CI: 58%-85%) of MAP cases in the entire internal test set as low risk, compared to mCTSI which identified 68% (95% CI: 53%-81%, p = 0.397). Importantly, neither method misclassified any SAP cases as low risk.

In the external test set, the AI model stratified 13% (95% CI: 10%-15%) of patients as high-risk and 23% (95% CI: 19%-27%) as low-risk. It successfully identified 73% (95% CI: 57%-88%) of SAP cases in the entire external test set as high-risk, compared to mCTSI which captured 55% (95% CI: 37%-71%, p = 0.035). The AI model demonstrated higher precision, with 1 SAP case per 2.8 high-risk patients (PPV: 0.357, 95% CI: 0.245–0.479), compared to 1 SAP case per 4.3 patients for mCTSI (PPV: 0.233, 95% CI: 0.143–0.333, p = 0.002). For low-risk triage, the AI model captured 40% (95% CI: 34%-46%) of MAP cases but mis-identified one SAP case as low-risk, whereas mCTSI identified 33% (95% CI: 28%-39%, p = 0.019) of MAP cases and mis-identified one SAP case as low-risk.

### Combining AI and mCTSI Predictions

We hypothesized that AI and mCTSI provide complementary strengths based on two key observations. First, Spearman’s correlation between their predictions was moderate on the validation set for both SAP (ρ = 0.602, 95% CI: 0.458–0.715) and MAP (ρ = 0.692, 95% CI: 0.582–0.777). Second, error analysis on the internal test set reveals that mCTSI and AI tend to make different errors (Figure 5). Among discordant cases, AI correctly predicted severity in all 5 instances where mCTSI overestimated (mCTSI ≥ 8), and in 8 of 9 cases where mCTSI underestimated severity (mCTSI < 2). In contrast, when mCTSI fell in the intermediate range (2 < mCTSI < 8), both methods showed similar accuracy. This finding suggests that mCTSI is more prone to error at extreme scores, where AI is more reliable. However, both methods perform similarly when the severity is moderate.

**Figure 5.**
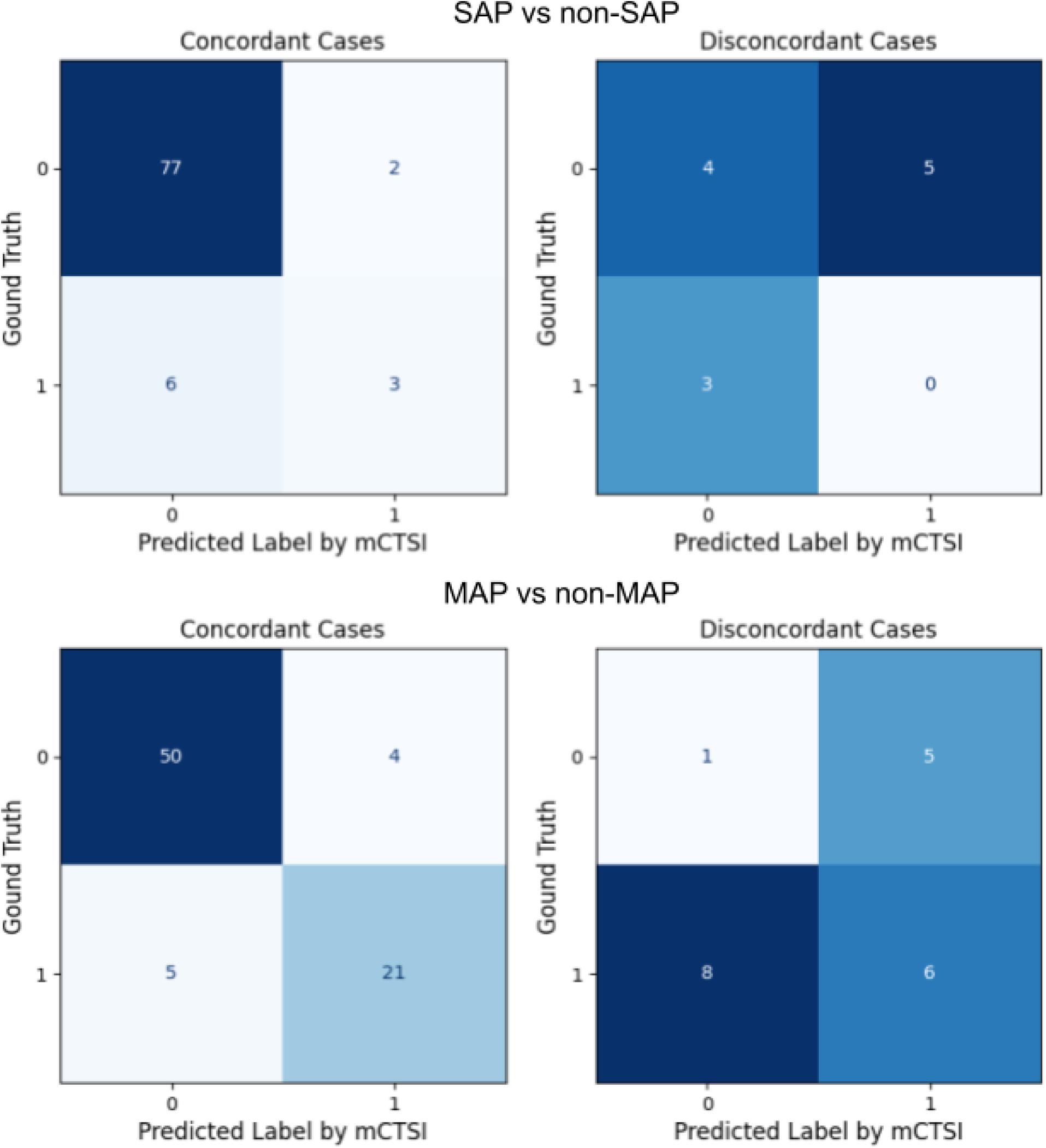
Error Analysis on the Internal Test Set. Confusion matrices are shown for cases where AI and mCTSI predictions are accordant (left column) and discordant (right column). The top row shows results for predicting future severe acute pancreatitis (SAP); the bottom row shows results for predicting mild acute pancreatitis (MAP). In discordant cases, mCTSI often errs when making extreme predictions, while AI tends to be correct in these scenarios.

Motivated by this observation, we developed a hybrid strategy that combines predictions from AI and mCTSI: for SAP prediction, we used AI scores alone when mCTSI ≥ 8 and averaged AI and mCTSI scores when mCTSI < 8; for MAP prediction, we used AI scores alone when mCTSI ≤ 2 and averaged the scores otherwise. On the internal test set, this approach yielded an AUROC of 0.891 (95% CI: 0.809–0.973) for SAP prediction, compared to 0.888 (95% CI: 0.800–0.960; p = 0.524) with AI alone, and 0.897 (95% CI: 0.834–0.960) for MAP prediction, compared to 0.888 (95% CI: 0.819–0.946; p = 0.358). On the external test set, the hybrid strategy achieved an AUROC of 0.895 (95% CI: 0.835–0.947) for SAP prediction, compared to 0.887 (95% CI: 0.825–0.941; p = 0.467) with AI alone. For MAP prediction, the hybrid model significantly improved AUROC from 0.858 (95% CI: 0.826–0.888) with AI alone to 0.874 (95% CI: 0.844–0.902; p = 0.027). These results demonstrate that the hybrid strategy improved predictive accuracy for both SAP and MAP across internal and external test sets, though the improvement did not reach statistical significance in most cases.

### Qualitative Case Study

We applied Gradient-weighted Class Activation Mapping (Grad-CAM)^54^ to generate heatmaps highlighting image regions the model considered important for its predictions. A board-certified abdominal radiologist with nine years of experience reviewed 20 SAP cases and found that the model tend to focused on imaging features clinically associated with severe AP, such as pancreatic parenchymal necrosis, acute peripancreatic fluid or necrotic collections, fat stranding, and ascites. Figure 6 and Supplementary Figure S7 illustrate three representative cases: one correctly identified only by the AI model (Figure 6), one correctly identified by both the AI model and mCTSI (Figure S7A), and one misclassified by both (Figure S7B). Notably, in cases where both the AI model and mCTSI failed to identify SAP, neither the Grad-CAM heatmaps nor the CT images showed clear radiologic markers to explain the severe clinical outcome.

**Figure 6.**
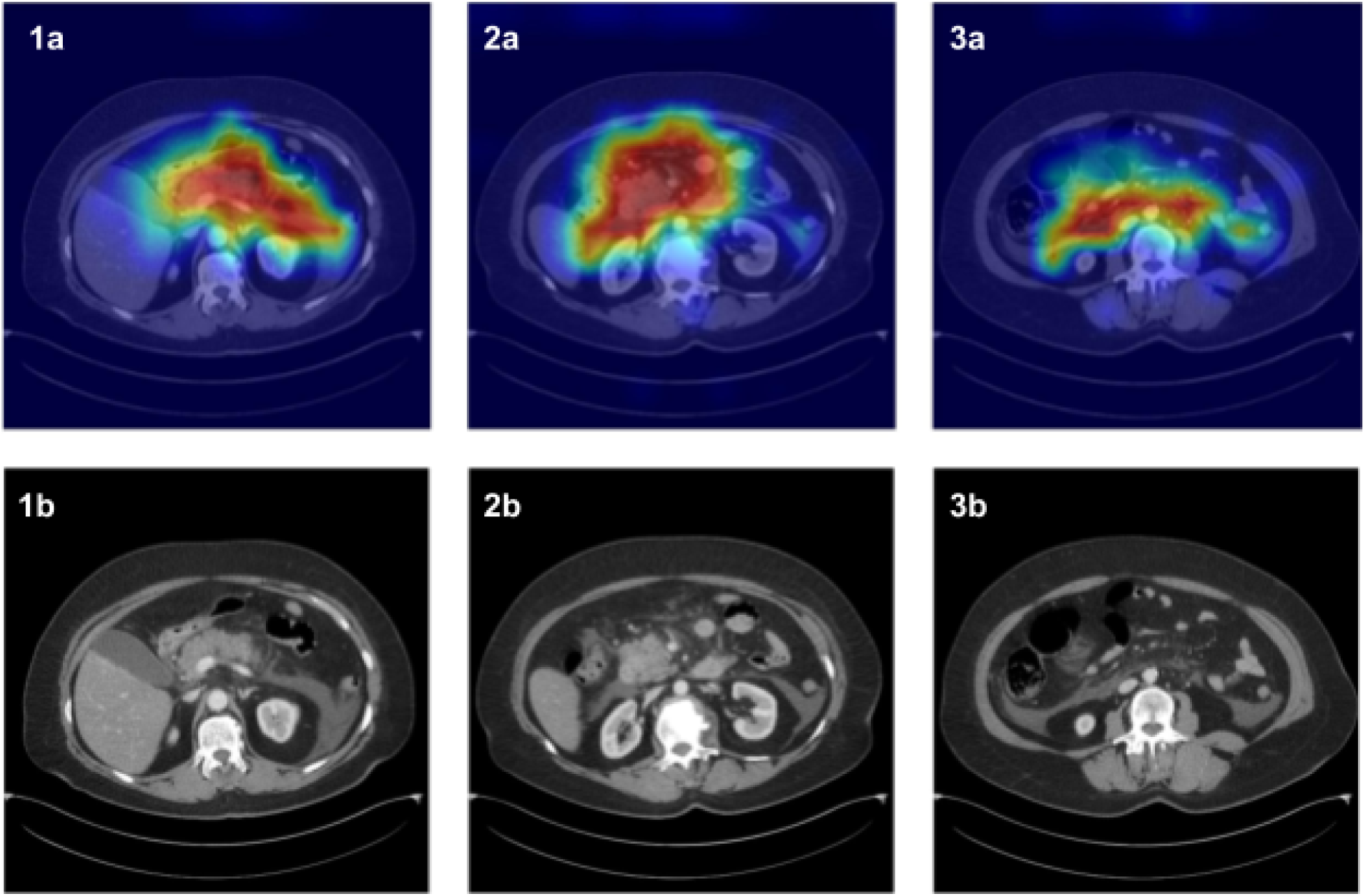
Heatmaps of a Severe Acute Pancreatitis Case from the External Test Set. Top row (1a, 2a, 3a): Grad-CAM heatmaps. Bottom row (1b, 2b, 3b): Corresponding axial portal-venous phase CT images. This case was correctly identified as severe by the AI model but was classified as moderately severe by mCTSI (score = 6). Each CT slice shows: 1. normal homogeneous enhancement of the pancreatic parenchyma, without pancreatic parenchymal necrosis (0 points); 2. acute peripancreatic fluid collection along the bilateral paracolic gutters (4 points) and 3. distant ascites more inferiorly in the abdomen (2 points). The three heatmaps indicate the model makes correct predictions highlighting the pancreatic inflammation, acute peripancreatic fluid collection and distant ascites respectively.

## Discussion

AP is a common but potentially life-threatening disease that requires early and accurate severity prediction to guide timely and effective clinical management. Existing severity prediction models used in research settings, such as the BISAP and mCTSI, have modest accuracy and often rely on laboratory results or imaging findings that may not be readily available at admission.^9,20,21^ These limitations reduce their practicality and contribute to their limited adoption in routine clinical practice.

To address these limitations, we developed an AI system that predicts AP severity from CECT scans acquired within 24 hours of admission. As CECT is routinely performed during the initial diagnostic work-up for patients presenting to the emergency room with abdominal pain in the U.S.,^55^ our model is able to provide risk prediction early in care without relying on inputs that may not be available yet. Moreover, our AI model demonstrated improved accuracy compared to mCTSI and BISAP. On the internal test set (n = 100), it achieved an AUROC of 0.888 for identifying patients who later developed SAP and 0.888 for those who developed MAP. On the external test set (n = 518), it achieved AUROCs of 0.887 for predicting SAP and 0.858 for predicting MAP, outperforming both mCTSI and BISAP. These results demonstrate the model’s potential to deliver accurate and automated risk assessment at early point of care.

One application of our AI model is at the time of admission, where it can be used to triage AP patients by identifying those suitable for early discharge and those who may require ICU-level care. To assess its clinical utility in this role, we conducted a retrospective simulation based on its severity predictions. This simulation-based validation approach is widely used in the literature to estimate AI models’ real-world impact.^56–58^ On the external test set, the AI model correctly identified 73% of all patients who eventually developed SAP as high-risk, with a precision of 35.7%, compared to mCTSI, which identified 55% of patients who developed SAP at 23.2% precision. Accurately identifying high-risk patients at admission is particularly valuable in low-resource settings where ICU resources are limited. Early recognition allows clinicians to allocate limited intensive monitoring to patients who truly need it and initiate timely escalation of care before clinical deterioration occurs. Conversely, on the internal test set, the AI model correctly identified 73% of low-risk patients who later developed only MAP without misclassifying any SAP cases, compared to 68% by mCTSI. Early identification of low-risk patients enables more efficient emergency department management by reducing unnecessary hospital admissions and expediting discharge for patients with mild AP.

Another application of our AI model is to serve as a second reader during interpretation of admission CECT scans, offering decision support to radiologists. Given the suboptimal inter-reader variability of mCTSI and its reduced sensitivity to early signs of SAP, our error analysis showed that AI can improve readers’ accuracy, especially when readers’ predictions are at the end of the spectrum (mCTSI ≤ 2 or ≥ 8). Building on this insight, we developed a simple heuristic to combine AI predictions with the radiologist’s mCTSI score. This approach achieved AUROCs of 0.895 for identifying high-risk patients and 0.874 for low-risk patients, compared to AI alone (0.888 and 0.858) and mCTSI alone (0.828 and 0.819). While these findings show the potential of AI to augment radiologist performance in early AP severity assessment, prospective clinical studies are needed to determine its true impact when integrated into real-world workflows.

Previous studies have investigated CT-based models for predicting AP severity, primarily using radiomic features^34,59,60^ or neural networks.^32,33,34^ Notably, Yin et al. proposed PrismSAP, a multi-modal risk model that combines clinical variables, radiomic features and CT images.^34^ This approach requires manual segmentation of worrisome features and manual selection of relevant CT slices, limiting scalability and introducing user variability. In contrast, our model does not require segmentation or manual slice selection at inference time, making it fully automated and more scalable for clinical use. Our work offers two key advancements over existing approaches.

First, most existing models were developed and tested using data from a single center.^32,33,59,60^ As a result, these models were not evaluated on truly independent external data from institutions that were not part of the training process, limiting their ability to demonstrate generalizability. In contrast, our model was trained exclusively on data from a large, diverse academic medical center in the U.S. and externally validated on an independent dataset from three medical centers in Hungary. While the internal and external test sets used the same inclusion criteria, the external CT scans varied in scanner models, vendors, acquisition parameters, and protocols. Despite these differences, our model demonstrated robust performance across both datasets, highlighting its strong generalizability across diverse patient populations and image acquisition protocols.

Second, unlike prior methods that rely solely on manually labeled severity levels, we utilized SSL methods to pretrain the model on large-scale unlabeled data. As demonstrated in our ablation study, SSL pre-training enables the model to learn generalizable imaging features, leading to improved accuracy in AP severity prediction. This strategy not only reduces reliance on manual annotations but also provides a scalable framework that is broadly applicable to other medical imaging tasks.^61,62^

Finally, while we aimed to compare our model against other AI-based approaches, most prior studies do not open-source their models and are trained on different datasets,^32,33,60^ making direct comparisons challenging. Reported performance metrics for AP severity prediction based on CT in existing studies range from 0.645 to 0.896 AUROC,^32–34^ which is comparable to the performance of our model. Unlike previous work, we open-source the weights and inference code of our model. This could enable transparent evaluation, reproducibility, and further development by the research community.

Despite its contribution, our study has several limitations. First, this was a retrospective study which may introduce selection bias, and would require prospective validation. Second, while CT scans are routinely obtained for patients presenting to the emergency room with abdominal pain in the U.S.,^31,55^ current guidelines recommend deferring CT imaging at admission if laboratory results and symptoms are sufficient to diagnose acute pancreatitis.^63^ This could limit the applicability of our approach. Third, our model does not incorporate clinical variables, including many easily obtainable laboratory markers such as BUN level, which could provide additional valuable prognostic information and further improve the performance of image-based models.

Finally, while our model demonstrated strong generalizability, its performance could benefit from a larger labeled training data. Expanding annotated datasets usually can enable the model to learn finer-grained imaging patterns while maintaining its robustness across diverse populations.

**Table 1.** Statistics of the Development Cohort and External Cohort. Summary of patient demographics, imaging findings, and acute pancreatitis severity classification based on the Revised Atlanta Classification criteria for both the Internal and External Datasets.

|  | Development and Internal Test Sets |  |  |  | External Test Set |
| --- | --- | --- | --- | --- | --- |
|  | Fine-tuning | Validation | Test | Total | Total |
| **Patients** | 528 | 75 | 100 | 703 |  |
| **CT Studies** | 550 | 75 | 100 | 725 |  |
| **Severity** |  |  |  |  |  |
| MAP | 245 (44.6%) | 31 (41.3%) | 40 (40.0%) | 316 (43.6%) | 276 (53.3%) |
| MSAP | 183 (33.3%) | 37 (49.3%) | 48 (48.0%) | 268 (37.0%) | 209 (40.4%) |
| SAP | 122 (22.2%) | 7 (9.3%) | 12 (12.0%) | 141 (19.5%) | 33 (6.4%) |
| **Demographics** |  |  |  |  |  |
| Gender, Female, n (%) | 222 (42.04%) | 32 (42.67%) | 41 (41%) | 295 (41.96%) | 212 (40.93%) |
| Age (mean, std) | 52.88 (18.16) | 51.53 (16.92) | 54.33 (18.01) | 52.94 (18.00) | 54.42 (14.83) |
| Height, cm (mean, std) | 1.69 (0.11) | 1.68 (0.11) | 1.67 (0.11) | 1.69 (0.11) | 1.70 (0.10) |
| Weight, kg (mean, std) | 82.28 (21.16) | 79.43 (19.03) | 80.62 (18.04) | 81.64 (20.42) | 82.39 (17.10) |
| **Imaging Findings** |  |  |  |  |  |
| Pleural fluid, n% | 13.79% | 12% | 15% | 13.76% | 7.24% |
| Peripancreatic fluid collection | 46.12% | 50.67% | 36% | 44.47% | 69.14% |
| Abdominal fluid | 44.05% | 36% | 45% | 42.79% | 47.18% |

## Data Availability

All data produced in the present study are not available for the public due to privacy protection.

## Acknowledgement

The authors thank Tedum Sampson for supporting the computing environment, Luoyao Chen and Harold Stern for supporting the data retrieval, and Benny Huang and Jason Tice for image extraction. This work was supported in part by grants from the National Science Foundation (1922658), and the Society of Abdominal Radiology GE Healthcare Research and Innovation Grant Award. This manuscript was edited for clarity using ChatGPT-4o.

## Conflict of interest statement

The authors declare no conflict of interest.

## Statement of whether data, analytic methods, and study materials will or will not be made available to other researchers

The data used in this study is not available for the public due to privacy protection. Upon completion of the review, the model’s inference code and trained weights will be released on GitHub under the MIT license at https://github.com/nyu-shenlab/ap-severity-evaluation.

## Notes

### Competing Interest Statement

The authors have declared no competing interest.

### Funding Statement

This study was funded in part by grants from the National Science Foundation (1922658), and the Society of Abdominal Radiology GE Healthcare Research and Innovation Grant Award.

### Author Declarations

Institutional Review Board of New York University Langone (IRB Protocol #I24-00008) waived ethical approval for this work.

