## Supplementary Materials for "Development and International Validation of a Deep Learning Model for Predicting Acute Pancreatitis Severity from CT Scans"

### S1. Image Acquisition, Preprocessing, and Labeling

#### S1.1 Image Acquisition Protocol

CT imaging at NYU Langone Health was performed using scanners from Siemens Healthineers, Canon Medical Systems, Philips Healthcare and GE HealthCare. Scans were obtained with slice thickness ranging from 0.2 to 5mm. Cube current and maximum CT energy varies based on patient size, height and BMI. Nonionic iodinated contrast media were injected using weight-based dosing. The agents used included iopamidol (Isovue 300 or Isovue 370; Bracco Imaging or Omnipaque 300; GE HealthCare) and iodixanol (Visipaque; GE HealthCare). The scan resolution is 512 by 512.

#### S1.2 Acute Pancreatitis Severity and mCTSI Labeling

Acute pancreatitis (AP) severity labeling followed a two-step manual process based on Revised Atlanta Classification. First, a radiology resident reviewed CT images after a training session, under the supervision of an attending radiologist, and recorded local complications. Second, a medical student, trained by a gastroenterology fellow, reviewed each patient's electronic health record (EHR) to document the presence and duration of organ failure, and assigned the final overall severity category.

Modified CTSI (mCTSI) scoring was independently evaluated by four senior radiology trainees (two PGY-5 and two PGY-6). To standardize scoring, all trainees were provided with training materials and deidentified example CT images from the collaborating external institution. Additionally, they participated in two virtual training sessions led by a radiologist from the external site. Each trainee then reviewed 100 unique CT scans, blinded to clinical information and RAC labels, and scored the degree of pancreatic inflammation, necrosis, and extrapancreatic complications according to mCTSI guidelines.

#### S1.3 Generating Segmentation Masks

We used MedSAM with the text prompt “pancreas” to generate pancreas segmentation masks. The input to MedSAM is a 2D CT slice along with a text prompt, and the output is a binary segmentation mask indicating whether each pixel belongs to the pancreas. To refine the masks, we applied morphological dilation and erosion to remove artifacts and ensure continuity of the segmented region.[64] For each CT series, we quantified pancreas size on each slice by counting the number of positive pixels in the mask. We then computed the total predicted pancreas volume across all 64-slice consecutive windows in the series and selected the window with the largest predicted volume for model training. Two examples of segmentation masks on axial slices are shown in Figure S1.

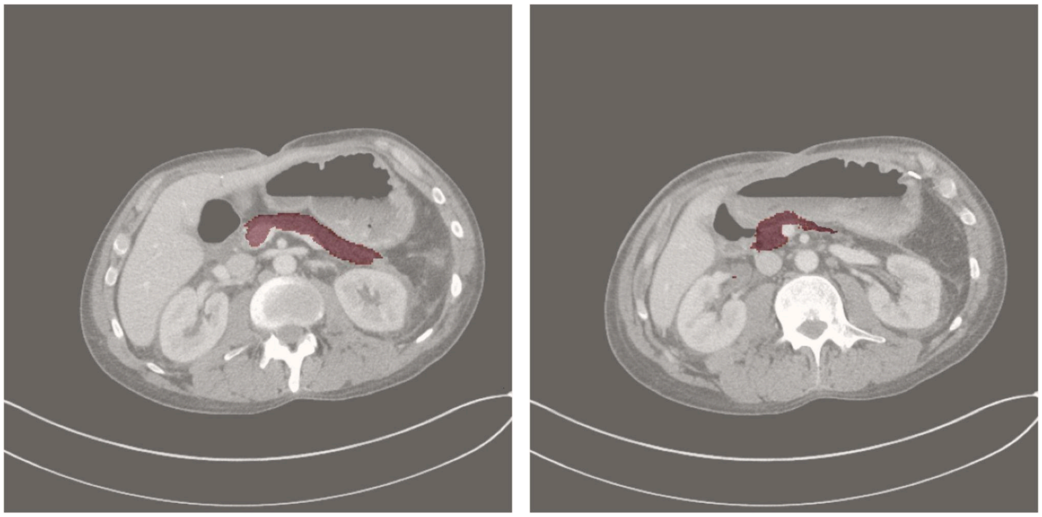

**Figure S1 Segmentation Masks Generated by MedSAM on Two Slices of a CT Study.** The generated masks cover pancreatic areas on both slices.

**S2. Extracting AP Severity Pseudo-Labels from Radiology Reports**

We manually reviewed 500 radiology reports to identify common text patterns in the findings section associated with each AP severity category: mild, moderately severe, and severe. For mild and moderately severe categories, we compiled a list of matching phrases, as shown in Table S2. If a report contained one of these phrases, we assigned the corresponding AP severity label to the associated CT scan as a pseudo-label. For the severe category, we searched for matches to the phrase “severe pancreatitis” across all reports. A radiologist then manually reviewed all matched cases to exclude false positives.

**Table S2. Matching Phrases in Radiology Reports Used for Automatic Pseudo-label Assignment.**

| Severity | Predefined Matching Phrases |
|---|---|
| Mild Acute Pancreatitis | "Mild acute pancreatitis",<br>"Findings consistent with mild acute pancreatitis",<br>"Mild acute pancreatitis as described above",<br>"Noncomplicated mild acute pancreatitis",<br>"compatible with mild acute pancreatitis",<br>"Interval resolution of a mild acute pancreatitis",<br>"Interval development of a mild acute pancreatitis",<br>"Mild acute pancreatitis of the distal pancreatic tail",<br>"Mildly edematous pancreas with mild surrounding stranding, increased since prior CT",<br>"Mild acute pancreatitis",<br>"Evidence of mild acute pancreatitis",<br>"Findings of mild acute pancreatitis", |
| Moderately Severe Acute Pancreatitis | "Moderate acute pancreatitis",<br>"Again seen is moderate acute pancreatitis",<br>"There is moderate acute pancreatitis", |

| | |
|---|---|
| | "This pattern indicates moderate acute pancreatitis.",<br>"consistent with moderate acute pancreatitis",<br>"consistent with a moderate acute pancreatitis",<br>"moderately severe acute pancreatitis",<br>"Moderate acute pancreatitis as described above",<br>"Changes of moderate acute pancreatitis",<br>"Findings compatible with moderate acute pancreatitis",<br>"Findings compatible with a moderate acute pancreatitis",<br>"Moderate acute pancreatitis without evidence of pancreatic necrosis",<br>"Moderate acute pancreatitis, appearing more pronounced when compared to prior CT",<br>"Moderate acute pancreatitis with slight interval improvement since prior CT",<br>"Interval of a moderate acute pancreatitis",<br>"Moderate acute pancreatitis, showing interval improvement",<br>"Moderate acute pancreatitis, with stable findings as detailed above",<br>"representing moderate acute pancreatitis" |

### S3. Model Training and Inference

#### S3.1 Preprocessing Input for Training and Inference

Let a CT study consist of $K$ axial series $\{S_1, S_2, ..., S_K\}$ and each series $S_i$ is a set of 2D slices. From each series $S_i$, we identify a subset of $N = 64$ pancreas-containing slices, denoted as:

$$X_{pan} = \{x_1, x_2, ..., x_N\},$$

To provide additional contextual information, we randomly sampled $M$ non-pancreas slices from the remaining slices in the same series, denoted as:

$$X_{other} = \{x'_1, x'_2, ..., x'_M\},$$

where $M$ is a hyperparameter. The final input for model training is defined as the union:

$$X = X_{pan} \cup X_{other}.$$

The selection of $X_{pan}$ was based on pancreas segmentation and was designed to cover the entire pancreas along with peri-pancreatic regions, which are critical for assessing inflammation, fat stranding, and fluid collections. The additional non-pancreas slices $X_{other}$ are included to provide contextual information, as inflammatory changes and fluid accumulation may also appear in adjacent or remote organs.

At each training epoch, one series was randomly selected from each CT study and preprocessed using this method. The random sampling of non-pancreas slices was repeated independently for each epoch to increase training diversity. During inference, the model generated a severity score for each available series in a CT study. The highest score across all series was used as the final severity prediction for that study.

#### S3.2 Training Details and Hyperparameter Tuning

We trained the model using a binary cross-entropy (BCE) loss to supervise the severity predictions task. To encourage meaningful and interpretable saliency maps, we added two regularization terms applied to the saliency maps $\hat{S}$. First, an L1 loss was used to promote sparsity, encouraging the model to highlight only the most relevant regions. Second, a Dice loss was applied to align the saliency maps $\hat{S}$ with pancreas segmentation masks $S$. The loss function for each task is defined as

$$L = BCE\left(y, \hat{y}\right) + \lambda_{dice} DICE\left(S, \hat{S}\right) + \lambda_{l1} L1\left(\hat{S}\right),$$

where $y$ is the ground-truth label, $\hat{y}$ is s the predicted score, $S$ is the pancreas segmentation mas, and $\hat{S}$ is the saliency map. The hyperparameters $\lambda_{dice}$ and $\lambda_{l1}$ control the strength of each regularization term. We trained the model to jointly predict both mild (MAP) and severe (SAP) AP. The total loss is defined as:

$$L_{total} = \alpha L_{SAP} + L_{MAP}$$

where each term corresponds to the loss for one prediction task and $\alpha$ is a hyperparameter that controls the contribution of the severe prediction loss to the overall training objective.

All models were trained for up to 60 epochs using the AdamW optimizer, with early stopping based on validation performance. A cosine learning rate scheduler was used, with a warm-up period over the first 2 epochs. The batch size was set to 2 during training and 1 during inference. To address class imbalance, CTs with SAP were upsampled twice in the training set. All models were trained using a single NVIDIA A100 GPU. Hyperparameters were tuned via random search, optimizing for AUROC on two classification tasks: MAP vs. non-MAP and SAP vs. non-SAP. The list of hyperparameters and their tuning ranges is provided in Table S3. Final performance was reported using an average ensemble of the top 5 models for each task, selected based on validation AUROC.

**Table S3. Hyperparameters and their corresponding ranges.**

| Hyperparameter | Ranges | Sampling Method |
|---|---|---|
| Learning rate | $[1e-5,\ 1e-4]$ | Uniform |
| Weight decay | $[1e-5,\ 1e-3]$ | Uniform |
| Top *k* | $[1e-1.3,\ 1e-0.5]$ | Uniform |
| Non pancreas slices $M$ | $[0,\ 16,\ 32,\ 48,\ 64]$ | Random Choice |
| L1 lambda $\lambda_{l1}$ | $[1e-5,\ 1e-4]$ | Uniform |
| DICE lambda $\lambda_{dice}$ | $[1e-2,\ 1e0.5]$ | Uniform |
| Pretrained weights | [*DINO, DINOv2, ImageNet*] | Random Choice |
| Loss weight for severe task $\alpha$ | $[1,\ 2]$ | Uniform |

#### S3.3 Data Augmentation

To improve model robustness and generalization, we applied data augmentations, independently to each slice, during training. Each CT slice was augmented using the following transformations:

- Random horizontal flip with a probability of 0.5.
- Random resized crop with a probability of 0.5, using a scale range of (0.8, 1.0) and an aspect ratio range of (0.8, 1.2).
- Random affine transformation with rotation degrees between -5° and 5°, translation up to 10% in both x and y directions, and scaling between 0.8× and 1.2×.
- Gaussian noise addition with a probability of 0.15.
- Random rotation up to ±15°.

**S4. Ablation Study Training Details and Results**

We adopted both DINO and DINOv2 for self-supervised learning, using the default hyperparameters as specified in their original papers. To select the best pretrained model, we performed linear probing, where a linear classifier was trained on frozen feature vectors extracted from the DINO teacher backbone. Each linear model was trained for 10 epochs using 80% of the fine-tuning set and evaluated on the remaining 20%. The pretrained model whose linear classifier achieved the highest macro AUROC across the SAP and MAP prediction tasks was selected for subsequent fine-tuning. To leverage SSL pretraining during fine-tuning, we initialize the encoder model with the weights selected from the SSL-pretrained model.

**Table S4. Ablation Study Results on Self-supervised Pretraining and Pseudo-labeled Data.** The baseline model was trained without self-supervised learning (SSL) or pseudo labels. Performance was evaluated using the area under the ROC curve (AUROC) on the external test set. Both techniques improved model performance for both the mild (MAP) vs. non-mild and severe (SAP) vs. non-severe tasks.

| Method | MAP vs non-MAP | SAP vs non-SAP |
|---|---|---|
| Baseline | 0.811 [0.773, 0.847] | 0.799 [0.717 0.871] |
| +Pseudo Labels | 0.834 [0.799, 0.868] | 0.845 [0.770, 0.907] |
| +SSL | 0.832 [0.796, 0.866] | 0.846 [0.773, 0.906] |
| +SSL & Pseudo Labels | 0.858 [0.826,0.888] | 0.887 [0.825, 0.941] |

**S5 Severity Prediction Performance on the Internal and External Test Sets.**

**Table S5.1. Performance Comparison of the AI Model, mCTSI, and the Hybrid Model on the Internal Test Set.** Evaluation metrics include area under the Receiver Operating Characteristic curve (AUROC), area under the precision-recall curve (AUPRC), sensitivity, specificity, positive predictive value (PPV), and negative predictive value (NPV) for identifying mild acute pancreatitis (MAP) and severe acute pancreatitis (SAP).

| | | AUROC | AUPRC | Sensitivity | Specificity | PPV | NPV |
|---|---|---|---|---|---|---|---|
| SAP vs non-SAP | mCTSI | 0.704 [0.551,0.838] | 0.205 [0.100,0.414] | 0.25 [0, 0.5] | 0.920 [0.859, 0.976] | 0.3 [0,0.600] | 0.9 [0.833,0.956] |
| | AI | 0.888 [0.800,0.960] | 0.566 [0.305,0.831] | 0.500 [0.200, 0.800] | 0.920 [0.859,0.976] | 0.462 [0.182, 0.750] | 0.931 [0.874,0.978] |
| | Hybrid | 0.891 [0.809, 0.973] | 0.578 [0.308, 0.840] | 0.500 [0.200, 0.786] | 0.955 [0.907, 0.989] | 0.600 [0.273, 0.909] | 0.933 [0.878, 0.978] |
| MAP vs non-MAP | mCTSI | 0.816 [0.733,0.890] | 0.688 [0.554,0.812] | 0.675 [0.525, 0.818] | 0.850 [0.755, 0.933] | 0.750 [0.600, 0.885] | 0,797 [0.694,0.892] |
| | AI | 0.888 [0.819,0.946] | 0.805 [0.672, 0.928] | 0.750 [0.610, 0.878] | 0.850 [0.754, 0.933] | 0.769 [0.629, 0.896] | 0.836 [0.737, 0.924] |
| | Hybrid | 0.897 [0.834, 0.960] | 0.810 [0.673, 0.933] | 0.525 [0.370, 0.682] | 0.933 [0.862, 0.985] | 0.840 [0.680, 0.964] | 0.747 [0.646, 0.841] |

**Table S5.2. Performance Comparison of the AI Model, BISAP, mCTSI and the Hybrid Model on the External Test Set.** Evaluation metrics include area under the Receiver Operating Characteristic curve (AUROC), area under the precision-recall curve (AUPRC), sensitivity, specificity, positive predictive value (PPV), and negative predictive value (NPV) for identifying mild acute pancreatitis (MAP) and severe acute pancreatitis (SAP).

| | | AUROC | AUPRC | Sensitivity | Specificity | PPV | NPV |
|---|---|---|---|---|---|---|---|
| SAP vs non-SAP | mCTSI | 0.828 [0.769,0.883] | 0.231 [0.135,0.363] | 0.545 [0.375, 0.718] | 0.878 [0.849, 0.907] | 0.234 [0.141, 0.333] | 0.966 [0.948, 0.982] |
| | BISAP | 0.693 [0.592,0.787] | 0.152 [0.082,0.272] | 0.242 [0.103, 0.4] | 0.957 [0.938, 0.974] | 0.276 [0.120, 0.444] | 0.949 [0.929, 0.967] |
| | AI | 0.887 [0.825, 0.941] | 0.442 [0.290, 0.639] | 0.727 [0.567, 0.875] | 0.878 [0.848, 0.907] | 0.289 [0.195, 0.388] | 0.979 [0.965, 0.991] |
| | Hybrid | 0.895 [0.835, 0.947] | 0.489 [0.335, 0.685] | 0.667 [0.5, 0.824] | 0.922 [0.897, 0.944] | 0.367 [0.25, 0.492] | 0.976 [0.961, 0.989] |
| MAP vs non-MAP | mCTSI | 0.819 [0.785,0.851] | 0.797 [0.752, 0.837] | 0.333 [0.279, 0.390] | 0.975 [0.954, 0.992] | 0.939 [0.888, 0.981] | 0.562 [0.515, 0.609] |

| | | | | | | | |
|---|---|---|---|---|---|---|---|
| | BISAP | 0.609 [0.564, 0.654] | 0.560 [0.548, 0.652] | 0.851 [0.809,0.891] | 0.285 [0.229,0.343] | 0.576 [0.529, 0.623] | 0.627 [0.535, 0.713] |
| | AI | 0.858 [0.826,0.888] | 0.866 [0.824, 0.905] | 0.380 [0.324, 0.439] | 0.975 [0.953, 0.992] | 0.946 [0.899, 0.983] | 0.580 [0.532,0.628] |
| | Hybrid | 0.874 [0.844, 0.902] | 0.886 [0.849, 0.919] | 0.232 [0.183, 0.282] | 0.996 [0.987, 1] | 0.985 [0.947, 1] | 0.532 [0.485, 0.576] |

### S6. Subcohort Analysis

**Table S6. Subcohort Analysis by Age and Gender.** Areas under the ROC curves (AUROCs) with 95% confidence intervals for different age and gender groups in the internal test set and external test set.

| | | | N | MAP vs non-MAP | SAP vs non-SAP |
|---|---|---|---|---|---|
| Internal Set | Age | > 55 | 51 | 0.862 [0.745, 0.959] | 0.886 [0.836, 1] |
| | | <= 55 | 49 | 0.906 [0812, 0.976] | 0.914 [0.796, 1] |
| | Gender | Male | 59 | 0.881 [0.782, 0.958] | 0.937 [0.836, 1] |
| | | Female | 41 | 0.911 [0.812, 0.981] | 0.887 [0.747, 0.986] |
| External Test Set | Age | > 55 | 358 | 0.867 [0.823, 0.908] | 0.870 [0.742, 0.920] |
| | | <= 55 | 260 | 0.851 [0.804, 0.894] | 0.930 [0.865, 0.980] |
| | Gender | Male | 306 | 0.853 [0.809, 0.893] | 0.840 [0.744, 0.920] |
| | | Female | 212 | 0.873 [0.824, 0.915] | 0.939 [0.856, 0.989] |

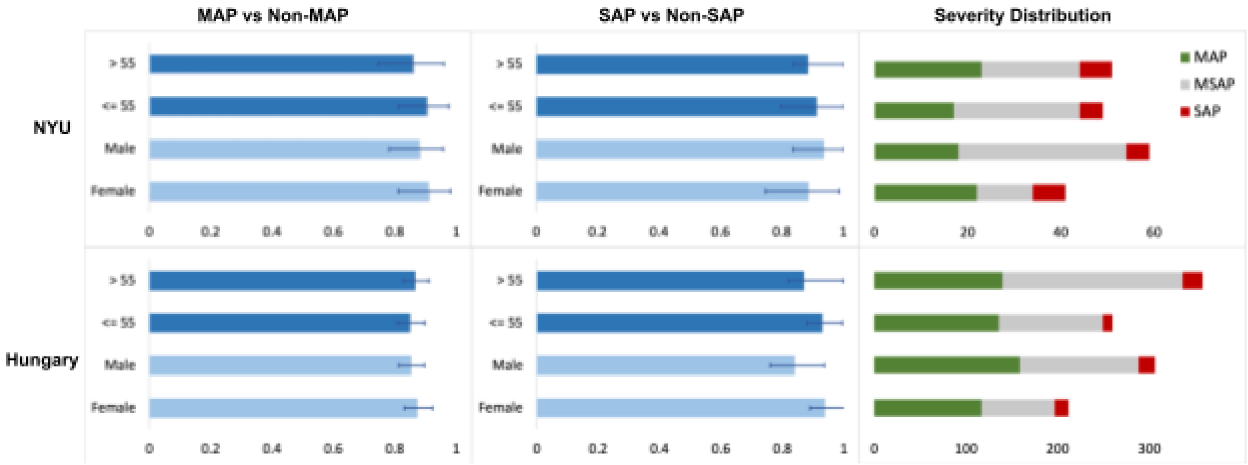


**Figure S6. Subgroup Analysis by Age and Gender on the Internal and External Test Sets.** From left to right: (1) AUROC with 95% confidence intervals for identifying mild acute pancreatitis (MAP), (2) AUROC with 95% confidence intervals for identifying severe acute pancreatitis (SAP), and (3) the distribution of mild, moderately severe, and severe cases across age and gender groups in both test sets.

### S7. Generating Heatmaps

Grad-CAM computes the gradients of the predicted class score with respect to the feature maps in the final layer of the image encoder. These weighted gradients are aggregated to produce an activation map, highlighting regions in the input image that most influence the model's prediction. In this work, Grad-CAM is used to generate slice-level heatmaps, which are then superimposed on the corresponding CT slices to indicate regions most relevant to the SAP prediction.

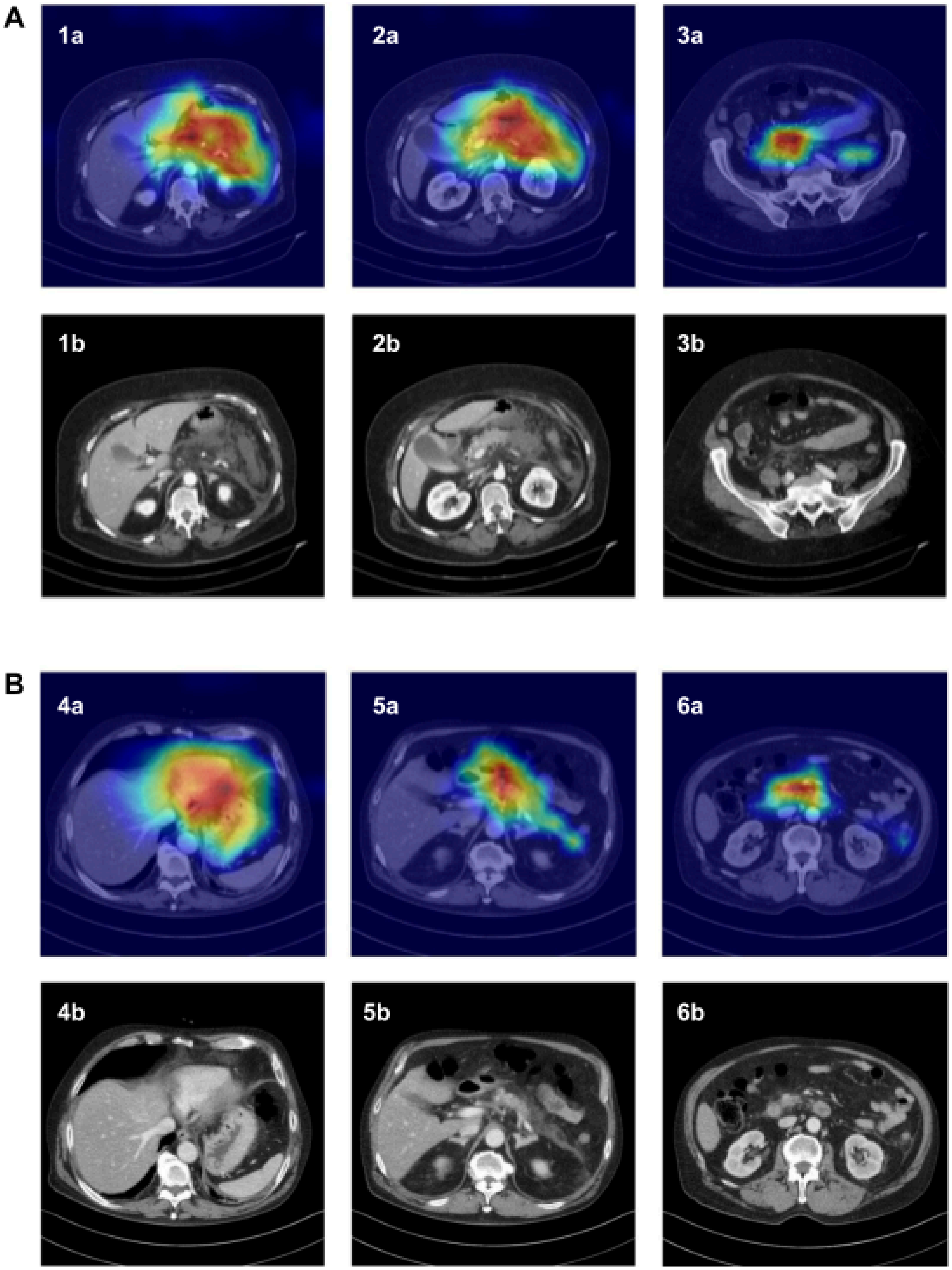


**Figure S7. Heatmaps of Two SAP Cases from the External Test Set.** Panel A shows a study that was correctly predicted as severe acute pancreatitis (SAP) by both the AI model and mCTSI (score = 10). Axial portal-venous phase CT images show >30% pancreatic parenchymal necrosis (4 points), peripancreatic acute necrotic collections (4 points), and ascites (2 points). Panel B shows a case misclassified by both the AI model and mCTSI (score = 6). CT findings include: no pancreatic parenchymal necrosis (0 points), pancreatic inflammation with peripancreatic fluid (4 points), and trace ascites inferiorly (2 points).
